# Uptake of service specific codes for the COVID oximetry @Home Pulse Oximetry service: an analysis of 57 million patients’ primary care records using OpenSAFELY

**DOI:** 10.1101/2025.04.07.25325394

**Authors:** Colm D Andrews, Christopher Wood, Louis Fisher, Helen J Curtis, William J Hulme, Amir Mehrkhar, Rebecca M Smith, David Evans, Tom Ward, Simon Davy, Peter Inglesby, Iain Dillingham, Steven Maude, Thomas O’Dwyer, Ben FC Butler-Cole, Lucy Bridges, Chris Bates, Jonathan Cockburn, Shaun O’Hanlon, Dima Avramov, Richard Jarvis, Alex J Walker, Ben Goldacre, The OpenSAFELY Collaborative, Brian MacKenna, Seb Bacon

**Affiliations:** Bennett Institute for Applied Data Science, Nuffield Department of Primary Care Health Sciences, Oxford University, Oxford, OX2 6GG; TPP, TPP House, 129 Low Lane, Horsforth, Leeds, LS18 5PX; EMIS,United Kingdom

## Abstract

**Background:** NHS England guidance was issued in November 2020 for use of COVID Oximetry @home (informally named CO@h) to detect early deterioration of patients with COVID-19 in primary and community care settings.

**Methods:** With the approval of NHS England we conducted a retrospective cohort study between 4^th^ October 2020 and 10^th^ July 2021, using the primary care records for 57 million people registered at an English general practice. We identified patients with pulse oximetry coding (either specific CO@H codes or non-specific COPD005 codes) and described their characteristics.

**Results:** We identified 18,473 individuals with a CO@h code, and 1,581,665 with a non-specific COPD005 code related to pulse oximetry. Recording of CO@h codes varied according to patient demographics, region and practice software system (58.3 per 100,000 in TPP vs 13.2 in EMIS).

**Conclusion:** Our study shows that whilst CO@h codes were used in GP records, use of less specific COPD005 codes instead of new SNOMED CT codes for CO@h may have persisted.

## Introduction

Severe hypoxaemia (low blood oxygen) without features of respiratory distress, known as silent hypoxia, is commonly seen in patients hospitalised with acute COVID-19 and is associated with worse patient outcomes^1^. Pulse oximeters are small portable medical devices that are attached to a person’s finger to measure blood oxygen levels. In November 2020, NHS England issued guidance for the use of remote pulse oximetry services as part of the COVID Oximetry @home service informally named by the NHS as CO@h^2^, to detect silent hypoxia in high risk patient groups with COVID-19 in primary and community care settings with the aim of enabling early escalation to hospital and reducing mortality^3,4^.

OpenSAFELY is a secure analytics platform for electronic health records (EHR) built by our group on behalf of NHS England to deliver urgent academic and operational research during the pandemic^5–9^. In England >99% of general practices use EHRs provided by EMIS or TPP. Using OpenSAFELY, researchers can run analyses across the full pseudonymised primary care records of patients registered at practices using TPP or EMIS, with patient-level linkage to various additional data sources within the TPP database.

NHS Digital released specific CO@h SNOMED CT codes^10^ that were made available in TPP from December 2020, and EMIS from January 2021 with the aim of supporting accurate reporting of this data, reducing the variability in data capture, and supporting clinical care of patients, research, policy making and healthcare planning. However, there are a number of other, less specific, codes available to record pulse oximetry, and it is possible that these alternative codes were used for some patients using the CO@h service, as they would be more familiar to clinicians. In particular, a broad set of pulse oximetry codes were previously (until 2019) financially incentivised in English primary care as part of a chronic obstructive pulmonary disease (COPD) Quality Outcomes Framework (QOF) indicator, known as COPD005^11^.

NHS Digital reported on the numbers of patients receiving CO@h until July 2022, using data gathered by clinical commissioning groups (CCG) or health service providers commissioned by CCGs^12^. They reported that 37,635 patients were onboarded to CO@h^13^. However there has been no data published on the recording of CO@h in primary care electronic health records.

We therefore set out to describe the use of SNOMED CT codes of both CO@h and COPD005 codes in primary care in England during the COVID-19 outbreak, broken down by patient demographics, clinical characteristics, and where possible their eligibility for CO@h.

## Methods

With the approval of NHS England we used OpenSAFELY to conduct a retrospective cohort study of pulse oximetry coding in the pseudonymised primary care records of general practices in England using either TPP or EMIS software, covering 57 million people.

Oximetry codes recorded between October 2020 and July 2021 were included, which was the latest date available at the time of analysis. All patients alive and registered at a TPP or EMIS practice as of 4 October 2020 were included. Patients whose recorded sex was not “male” or “female” were excluded.

Our outcomes were (1) any recorded CO@h SNOMED CT code and (2) any recorded COPD005 code^11,14^ (Box 1). We report the total number of patients with at least one of these outcomes, and the proportion per 100,000 patients including 95% confidence intervals. We also report the rate of coding over time using the first instance of a relevant code per patient.

Counts and rates of recorded events were stratified by various characteristics, ascertained as at 4 October 2020:

- Age, categorised as 0-17, 18-24, 25-34, 35-44, 45-54, 55-69, 70-79, 80+.
- Sex, using categories “male” and “female”, matching the ONS recorded categories;.
- Deprivation, measured by the 2019 Index of Multiple Deprivation (IMD) derived from the patient’s postcode at lower super output area level. IMD was divided by quintile, with 1 representing the most deprived areas and 5 representing least deprived areas. Where a patient’s postcode cannot be determined the IMD is recorded as unknown.
- Ethnicity, self-reported ethnicity categorised as Asian, Black, Mixed, White, Other, Unknown.
- Region, defined as the Nomenclature of Territorial Units for Statistics (NUTS 1) region derived from the patient’s practice’s postcode.
- Eligibility for pulse oximetry (in OpenSAFELY-TPP only), based on CO@h guidance at the time of the study^15^ (Box 2). Eligibility for pulse oximetry could not be ascertained from EMIS as OpenSAFELY-EMIS did not have linked COVID-19 infection data from the national Second Generation Surveillance System (SGSS) at the time of the study.

Counts and rates within each subgroup were reported by GP system (TPP or EMIS) and overall.

### Data Source

All data were linked, stored and analysed securely using the OpenSAFELY platform, https://www.opensafely.org/, as part of the NHS England OpenSAFELY COVID-19 service. Data include pseudonymised data such as coded diagnoses, medications and physiological parameters. No free text data are included. Detailed pseudonymised patient data is potentially re-identifiable and therefore not shared. All code is shared openly for review and re-use under MIT open license (https://github.com/opensafely/SRO-PULSE-OXIMETRY-UPD).

### Software and Reproducibility

Data management was performed using Python 3.8, with analysis carried out using Python. Code for data management and analysis, as well as codelists, are archived onlineData management was performed using Python 3.8, with analysis carried out using Python. Code for data management and analysis, as well as codelists, are archived online (https://github.com/opensafely/sro-pulse-oximetry-upd). All code for the OpenSAFELY platform for data management, analysis and secure code execution is shared for review and re-use under open licences at <u>github.com/opensafely-core</u>.

This was an analysis delivered using federated analysis through the OpenSAFELY platform. A federated analysis involves carrying out patient level analysis in multiple secure datasets, then later combining them: codelists and code for data management and data analysis were specified once using the OpenSAFELY tools; then transmitted securely from the OpenSAFELY jobs server to the OpenSAFELY-TPP platform within TPP’s secure environment, and separately to the OpenSAFELY-EMIS platform within EMIS’s secure environment, where they were each executed separately against local patient data; summary results were then reviewed for disclosiveness, released, and combined for the final outputs. All code for the OpenSAFELY platform for data management, analysis and secure code execution is shared for review and re-use under open licences on GitHub: https://github.com/OpenSAFELY.

#### Patient and Public Involvement and Engagement (PPIE)

OpenSAFELY has involved patients and the public in various ways: we developed a public website that provides a detailed description of the platform in language suitable for a lay audience (https://opensafely.org); we have participated in two citizen juries exploring public trust in OpenSAFELY; we have co-developed an explainer video (https://www.opensafely.org/about/); we have patient representation who are experts by experience on our OpenSAFELY Oversight Board; we have partnered with Understanding Patient Data to produce lay explainers on the importance of large datasets for research; we have presented at various online public engagement events to key communities (e.g., Healthcare Excellence Through Technology; Faculty of Clinical Informatics annual conference; NHS Assembly; HDRUK symposium); and more. To ensure the patient voice is represented, we are working closely to decide on language choices with appropriate medical research charities (e.g., Association of Medical Research Charities). We will share information and interpretation of our findings through press releases, social media channels, and plain language summaries.

## Results

The combined cohort included 57.4m people alive and registered as at 4 October 2020, 24.2m in TPP practices and 33.2m in EMIS practices. Of these 50% were female; 62% were white and 24% had no recorded ethnicity.

We identified 18,473 patients with at least one CO@h code (Table 1) and 1,581,665 with a COPD005 code (Table 2). There was a substantially higher rate of CO@h code use in TPP (58.3 per 100,000) compared to EMIS (13.2).

**Table 1:**
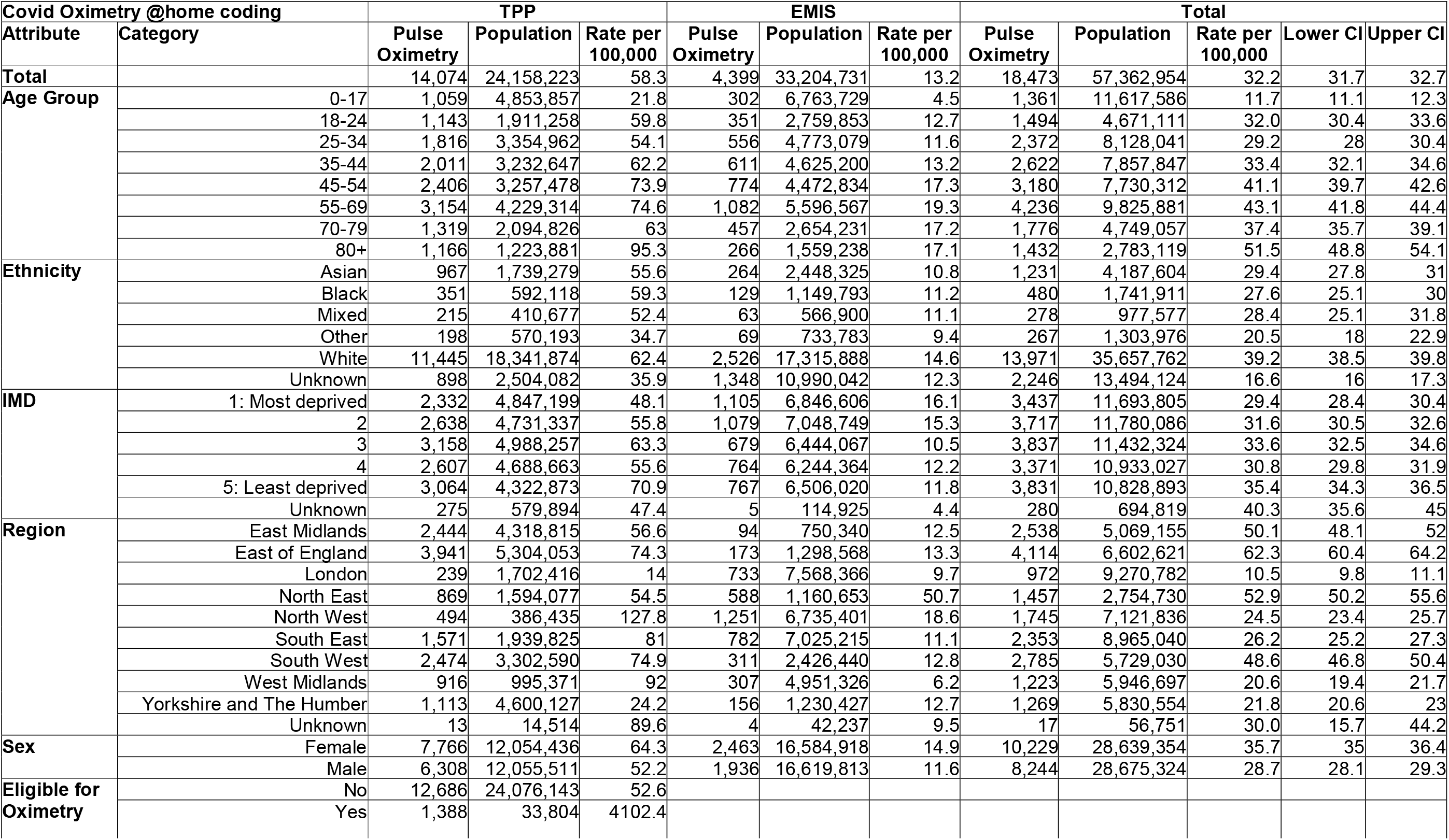
Counts and rates of patients with Covid Oximetry @home coding between 2020-10-04 and 2021-07-10, stratified by demographic variable. “Eligible for Oximetry” indicates patients diagnosed with COVID-19 and symptomatic and either aged 65 years or older or under 65 years and at higher risk from COVID-19, or where clinical judgement applies considering individual risk factors such as pregnancy, learning disability, caring responsibilities and/or deprivation. It was only possible to ascertain eligibility for oximetry in TPP. Due to the geographic distribution of TPP and EMIS practices, regions may not represent the whole region^7^. IMD= Index of Multiple Deprivation

**Table 2:**
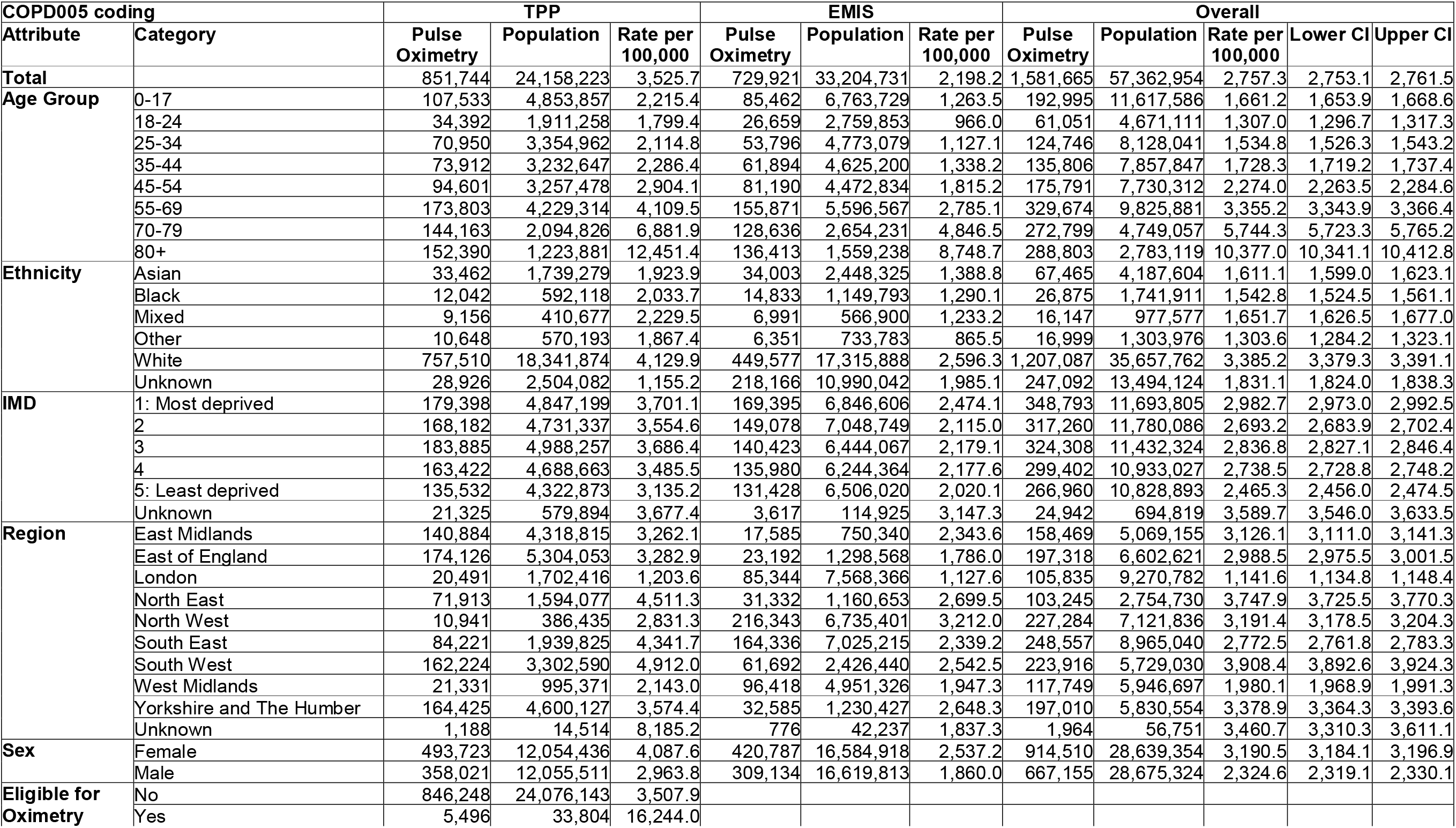
Counts and rates of patients with COPD005 coding between 2020-10-04 and 2021-07-10, stratified by demographic variable. “Eligible for Oximetry” indicates patients diagnosed with COVID-19 and symptomatic and either aged 65 years or older or under 65 years and at higher risk from COVID-19, or where clinical judgement applies considering individual risk factors such as pregnancy, learning disability, caring responsibilities and/or deprivation. It was only possible to ascertain eligibility for oximetry in TPP.

There was an initial increase in CO@h code use in both TPP and EMIS in January to February 2021. Usage gradually decreased until a second increase in usage in mid-late June 2021. Use of COPD005 codes was much higher and more variable but showed a gradual increase from early to mid 2021 (Figure 1).

**Figure 1:**
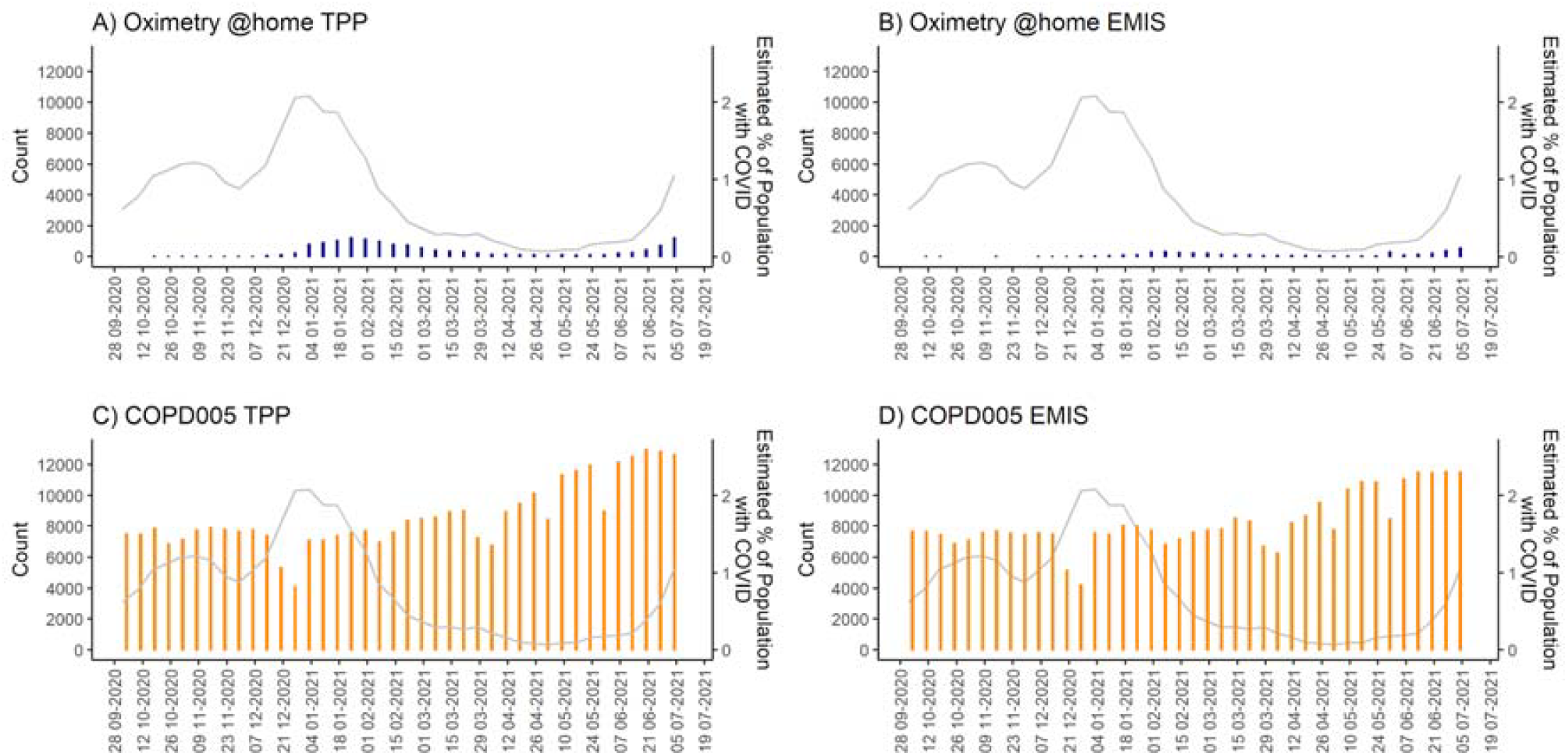
Barplot showing number of patients with Oximetry@home codes and COPD005 codes per week between 2020-10-04 and 2021-07-10, stratified by the electronic health record provider of the practice (TPP or EMIS) with line graph of official reported estimates of the percentage of the population testing positive for COVID-19 (ONS Coronavirus (COVID-19) Infection Survey). Guidance on COVID Oximetry @home was rolled out nationally in November 2020.

Table 2 shows demographic and clinical breakdowns of CO@h use. There was a higher rate of CO@h code use in women (35.7 per 100,000) than in men (28.7). The highest rate of code use was in white patients (39.2) with “unknown” ethnicity being lowest (16.6), 27.6 for Black patients and 29.4 for South Asian patients. There was substantial regional variation, ranging from 10.5 in London to 62.3 in the East of England.

Over half of TPP practices did not use any CO@h codes (51.6%) whereas only 0.5% of TPP practices did not use any COPD005 codes during the study period. Over two thirds of EMIS practices did not use any CO@h codes (79.3%) and 1.2% of EMIS practices did not use any COPD005 codes.(Figure 2). Among the CO@h codes, the most commonly used was “*Telehealth pulse oximetry monitoring started*” for both the general population (n= 6,946, 20%) and those eligible for pulse oximetry (n=1,073, 27.9%). Among the COPD005 codes, “*Peripheral oxygen saturation*” was the most commonly used code for both the general population (n= 1,696,810, 85%) and those eligible for pulse oximetry (n=6,582, 68.5%) (Tables 3 and 4). CO@h codes also included codes indicative of pulse oximetry not being suitable, e.g “*not appropriate*” (n= 6,545,18%) and “*declined*” (n= 1,823, 5%) (Table 3).

**Table 3:** Total use of each individual Pulse Oximetry related code between 2020-10-04 and 2021-07-10, grouped into CO@h codes and COPD005 codes. This counts all coded events, including where patients have been coded more than once.

| Code | Term | TPP |  | EMIS |  | Overall |  |
| --- | --- | --- | --- | --- | --- | --- | --- |
|  |  | N | % | N | % | N | % |
| CO@h |  |  |  |  |  |  |  |
| 1325191000000108 | Telehealth pulse oximetry monitoring started | 5,898 | 20.0 | 1,048 | 17.2 | 6,946 | 19.6 |
| 1325201000000105 | Telehealth pulse oximetry monitoring ended | 2,867 | 9.7 | 518 | 8.5 | 3,385 | 9.5 |
| 1325211000000107 | Provision of pulse oximeter | 4,843 | 16.4 | 1,073 | 17.6 | 5,916 | 16.7 |
| 1325221000000101 | Telehealth pulse oximetry monitoring not appropriate | 5,445 | 18.5 | 1,100 | 18.0 | 6,545 | 18.4 |
| 1325241000000108 | Telehealth pulse oximetry monitoring declined | 1,459 | 5.0 | 364 | 6.0 | 1,823 | 5.1 |
| 1325251000000106 | Referral to telehealth pulse oximetry monitoring service | 2,993 | 10.2 | 632 | 10.4 | 3,625 | 10.2 |
| 1325261000000109 | Referral by telehealth pulse oximetry monitoring service | 149 | 0.5 | 53 | 0.9 | 202 | 0.6 |
| 1325270000000000 | Discharge from telehealth pulse oximetry monitoring service | 1,273 | 4.3 | 448 | 7.3 | 1,721 | 4.8 |
| 1325281000000100 | Discussion about telehealth pulse oximetry monitoring | 4,357 | 14.8 | 469 | 7.7 | 4,826 | 13.6 |
| 1325681000000102 | Has access to pulse oximeter | 137 | 0.5 | 356 | 5.8 | 493 | 1.4 |
| 1325691000000100 | Oxygen saturation at periphery unknown | 12 | 0.0 | 35 | 0.6 | 47 | 0.1 |
| 1325701000000100 | Oxygen saturation at periphery equivocal | 29 | 0.1 |  |  |  |  |
| COPD005 |  |  |  |  |  |  |  |
| 252465000 | Pulse oximetry | 50,073 | 91.4 | 32,605 | 3.6 | 82,678 | 4.1 |
| 431314004 | Peripheral oxygen saturation | 987,903 | 4.6 | 708,907 | 77.7 | 1,696,810 | 85.1 |
| 866661000000106 | Peripheral blood oxygen saturation on room air at rest | 28,225 | 2.6 | 60,042 | 6.6 | 88,297 | 4.4 |
| 866701000000100 | Peripheral blood oxygen saturation on supplemental oxygen at rest | 408 | 0.0 | 1,249 | 0.1 | 1,657 | 0.1 |
| 927981000000106 | Baseline oxygen saturation at periphery | 14,608 | 1.4 | 109,924 | 12.0 | 124,532 | 6.2 |

**Table 4:** Total use of each individual Pulse Oximetry related code in those “Eligible for Oximetry” in OpenSAFELY-TPP between 2020-10-04 and 2021-07-10: “Eligible for Oximetry” indicates patients diagnosed with COVID-19 and symptomatic and either aged 65 years or older or under 65 years and at higher risk from COVID-19, or where clinical judgement applies considering individual risk factors such as pregnancy, learning disability, caring responsibilities and/or deprivation. It was only possible to ascertain eligibility for oximetry in TPP. This counts all coded events, including where patients have been coded more than once.

| Code | Term | Total records | % |
| --- | --- | --- | --- |
| **CO@h** |  |  |  |
| **1325191000000108** | Telehealth pulse oximetry monitoring started | 1,073 | 27.9 |
| **1325201000000105** | Telehealth pulse oximetry monitoring ended | 349 | 9.1 |
| **1325211000000107** | Provision of pulse oximeter | 663 | 17.3 |
| **1325221000000101** | Telehealth pulse oximetry monitoring not appropriate | 235 | 6.1 |
| **1325241000000108** | Telehealth pulse oximetry monitoring declined | 148 | 3.9 |
| **1325251000000106** | Referral to telehealth pulse oximetry monitoring service | 439 | 11.4 |
| **1325261000000109** | Referral by telehealth pulse oximetry monitoring service | 36 | 0.9 |
| **1325271000000102** | Discharge from telehealth pulse oximetry monitoring service | 165 | 4.3 |
| **1325281000000100** | Discussion about telehealth pulse oximetry monitoring | 726 | 18.9 |
| **1325681000000102** | Has access to pulse oximeter | 9 | 0.2 |
| **1325691000000100** | Oxygen saturation at periphery unknown | 0.0 | 0.0 |
| **1325701000000100** | Oxygen saturation at periphery equivocal | 0.0 | 0.0 |
| **COPD005** |  |  |  |
| **431314004** | Peripheral oxygen saturation | 6,582 | 68.5 |
| **252465000** | Pulse oximetry | 872 | 9.1 |
| **866661000000106** | Peripheral blood oxygen saturation on room air at rest | 1,506 | 15.7 |
| **866701000000100** | Peripheral blood oxygen saturation on supplemental oxygen at rest | 8 | 0.1 |
| **927981000000106** | Baseline oxygen saturation at periphery | 635 | 6.6 |

**Figure 2:**
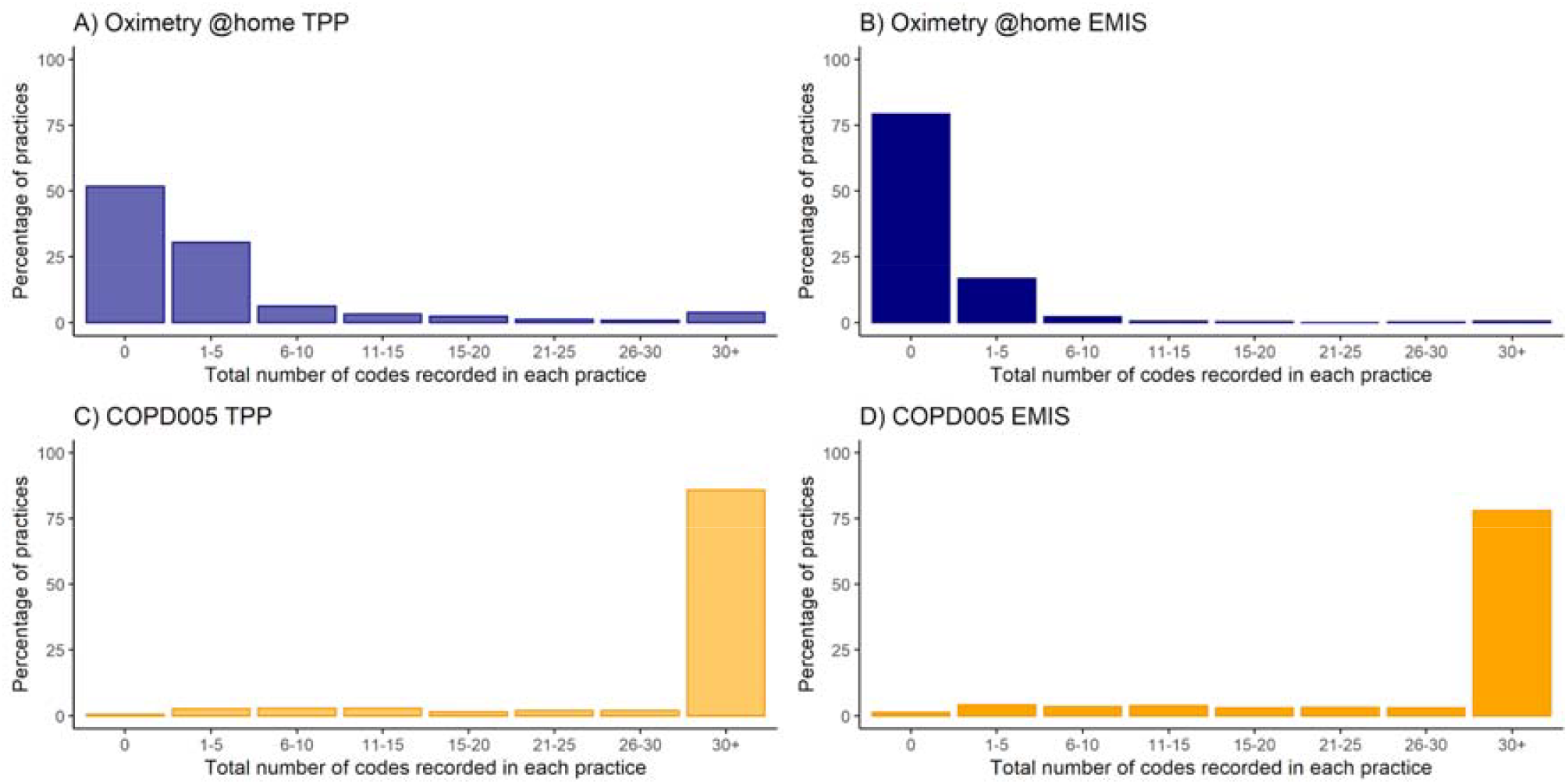
Volume of codes recorded per practice with at least one Covid Oximetry @home or COPD005 code between 4th October 2020 and 10th July 2021, stratified by the electronic health record provider of the practice (TPP or EMIS).

### Eligibility

From the TPP cohort, we ascertained 33,804 patients as eligible for CO@h (Table 1), of whom 1,388 (4102.4 per 100,000) had a CO@h code and 5,496 (16,244 per 100,000) had a COPD005 code. There was a substantially higher rate of recording of both COPD005 and CO@h in the eligible cohort (16,244 and 4,102 per 100,000 respectively) compared to the rest of the TPP cohort (3,507.9 and 52.6) (Table 1-2). Those eligible for CO@h accounted for 9.9% of the CO@h code use and 0.6% of the COPD005 code use.

## Discussion

We identified 18,473 patients in England with at least one CO@h code between 4^th^ October 2020 and 10^th^ July 2021, indicating that they received at-home monitoring for COVID-19-related hypoxia, and 1,581,665 with a COPD005 code which may be an indicator of similar treatment. However, the very small overlap between eligibility for CO@h and COPD005 code use indicates the vast majority of COPD005 use was likely unrelated to CO@h. The rate of CO@h code usage increased with the rise in COVID-19 cases in both January and June 2021^16^. There was a rise in use of COPD005 codes from March 2021, but with no marked increase in January. There was a surprisingly small overlap with treatment eligibility, with less than 10% of patients with a CO@h code being identified as eligible for this treatment.

### Interpretation

The COPD005 codes were historically used for an unrelated financially incentivised indicator. Individually the codes also represent aspects of a procedure (pulse oximetry) frequently carried out in primary care. It is possible that GPs have been using these COPD005 pulse oximetry codes for CO@h, or use may reflect a continued use for COPD reviews or standard clinical practice.There was a large increase in usage of the COPD005 codes during our study period but there wasn’t a strong overlap with patients eligible for CO@h. Although just 10% of patients with a CO@h code were identified as eligible for this treatment 4% of the eligible population had a CO@h code compared to 0.05% of the ineligible population.

We found large variations in the rate of use of CO@h codes between different geographic regions, practices, IMD quintiles, ethnicity, and sex. This may reflect differences in disease prevalence or service provision, a lack of recording, or the use of COPD005 codes which are not specific to the CO@h service to denote pulse oximetry. There was a substantially higher rate of recording of CO@h in practices that use TPP compared to those that use EMIS. However CO@h codes were made available earlier in TPP than EMIS^17^, impacting uptake earlier in the study period due to unavailability of the codes in EMIS. The difference may also be explained by the geographic clustering of EHR systems^18^ and the clinical caseload being different in those areas.

### Strengths and limitations

The OpenSAFELY platform has enabled analysis to an unprecedented scale and completeness of data. This is a study of coding and not service provision therefore further work would be required to distinguish between COPD005 codes used for CO@h monitoring and other uses. Patients were eligible for CO@h if they were diagnosed with COVID-19 and symptomatic and either aged 65 years or older or under 65 years and at higher risk from COVID-19, or where clinical judgement applies considering individual risk factors such as pregnancy, learning disability, caring responsibilities and/or deprivation. Our coding of eligibility does not take individual clinical judgement into account and as such could underestimate the true number of eligible people in some groups or misclassify some people. Our study used patient age as of October 2020 so may exclude patients who became 65 during the study period. In addition codes to indicate symptomatic status may be missing from GP records. In some cases Pulse oximetry services are managed by community teams who may not be able to add codes directly to GP records^19^. We, therefore, may not have access to all uses of CO@h codes.

### Findings in context

A study from a regional NHS organisation found that CO@h improved patient outcomes, reducing the odds of longer length hospital stays and mortality^4^. NHS England have supported pulse oximetry in CO@h to fulfil the objective of ensuring more timely hospital treatment for those who need it^15^. Considerable regional variation in uptake of CO@h has been reported^20^. Data submitted by regional teams to NHS Digital (NHSD) Covid Oximeter Weekly Report showed that 26,655 patients were onboarded to CO@h during our study period^13^, whereas we found 18,473 patients with a CO@h code (including those who may have declined). Several factors are likely to contribute to this discrepancy: regional teams submit the information to NHSD via a different unvalidated mechanism,^19^ other local reporting arrangements are in place^21^, delays in implementation of the relevant codes in EHRs may have prevented GPs from coding patients and pulse oximetry services are managed in some cases by community teams who may not be able to add codes directly to GP records. The difference is also likely exacerbated by the use of COPD005 codes instead of the CO@h codes.

Our data allowed us to assess eligibility in TPP. We found a marked increase in the rate of usage of both the CO@h SNOMED CT codes and COPD005 codes in eligible patients compared to ineligible patients. The NHSD Covid Oximeter Weekly Report did not include the rate of onboarding by eligibility.

### Policy Implications

In order to inform services such as CO@h, high quality data on onboarded patients and their outcomes should be captured. Our counts of SNOMED CT codes being lower than NHSD reports indicates that the CO@h SNOMED CT codes were not always applied in GP records for patients onboarded to this service. Manual data collection has drawbacks; it is time-consuming and does not easily permit linkage to other data such as patient outcomes. Through the OpenSAFELY platform, data on the use of CO@h SNOMED CT codes can be linked to externally linked datasets in order to perform a more detailed impact assessment of the CO@H service. Usage of dedicated SNOMED CT codes and/or standardised software templates could be encouraged to permit automated reporting and further analysis.

There may have been incomplete dissemination of information relating to the new codes specific for this service leading to usage of alternative COPD005 codes in some cases. Discrepancies between demographic groups may require further investigation, e.g. usage appeared lower in non-white ethnicities despite evidence of worse COVID outcomes; however it is possible this may be explained by geographic/software differences^7^.

## Conclusions

Our study shows that whilst CO@h codes were used in GP records, the use of less specific COPD005 codes instead of new SNOMED CT codes for CO@h may have persisted. Total population data from NHS general practice, accessed via OpenSAFELY, can facilitate new insights into service delivery.

## Data Availability

Access to the underlying identifiable and potentially re-identifiable pseudonymised electronic health record data is tightly governed by various legislative and regulatory frameworks, and restricted by best practice. The data in the NHS England OpenSAFELY COVID-19 service is drawn from General Practice data across England where EMIS and TPP are the data processors.
EMIS and TPP developers initiate an automated process to create pseudonymised records in the core OpenSAFELY database, which are copies of key structured data tables in the identifiable records. These pseudonymised records are linked onto key external data resources that have also been pseudonymised via SHA-512 one-way hashing of NHS numbers using a shared salt. University of Oxford, Bennett Institute for Applied Data Science developers and PIs, who hold contracts with NHS England, have access to the OpenSAFELY pseudonymised data tables to develop the OpenSAFELY tools.
These tools in turn enable researchers with OpenSAFELY data access agreements to write and execute code for data management and data analysis without direct access to the underlying raw pseudonymised patient data, and to review the outputs of this code. All code for the full data management pipeline - from raw data to completed results for this analysis - and for the OpenSAFELY platform as a whole is available for review at github.com/OpenSAFELY.

## Administrative

## Acknowledgements

We are very grateful for all the support received from the EMIS and TPP Technical Operations teams throughout this work, and for generous assistance from the information governance and database teams at NHS England and the NHS England Transformation Directorate.

## Funding

The OpenSAFELY platform is principally funded by grants from:

NHS England [2023-2025]; The Wellcome Trust (222097/Z/20/Z) [2020-2024]; MRC (MR/V015737/1) [2020-2021].

Additional contributions to OpenSAFELY have been funded by grants from:

MRC via the National Core Study programme, Longitudinal Health and Wellbeing strand (MC_PC_20030, MC_PC_20059) [2020-2022] and the Data and Connectivity strand (MC_PC_20058) [2021-2022];NIHR and MRC via the CONVALESCENCE programme (COV-LT-0009, MC_PC_20051) [2021-2024]; NHS England via the Primary Care Medicines Analytics Unit [2021-2024].

The views expressed are those of the authors and not necessarily those of the NIHR, NHS England, UK Health Security Agency (UKHSA), the Department of Health and Social Care, or other funders. Funders had no role in the study design, collection, analysis, and interpretation of data; in the writing of the report; and in the decision to submit the article for publication.

## Patient and Public Involvement and Engagement

OpenSAFELY has involved patients and the public in various ways: we developed a public website that provides a detailed description of the platform in language suitable for a lay audience (https://opensafely.org); we have participated in two citizen juries exploring public trust in OpenSAFELY; we have co-developed an explainer video (https://www.opensafely.org/about/); we have patient representation who are experts by experience on our OpenSAFELY Oversight Board; we have partnered with Understanding Patient Data to produce lay explainers on the importance of large datasets for research; we have presented at various online public engagement events to key communities (e.g., Healthcare Excellence Through Technology; Faculty of Clinical Informatics annual conference; NHS Assembly; HDRUK symposium); and more. To ensure the patient voice is represented, we are working closely to decide on language choices with appropriate medical research charities (e.g., Association of Medical Research Charities). We will share information and interpretation of our findings through press releases, social media channels, and plain language summaries.OpenSAFELY has developed a publicly available website https://www.opensafely.org/ through which they invite any patient or member of the public to make contact regarding the broader OpenSAFELY project.

## Information governance and ethical approval

NHS England is the data controller of the NHS England OpenSAFELY COVID-19 Service; EMIS and TPP are the data processors; all study authors using OpenSAFELY have the approval of NHS England^22^. This implementation of OpenSAFELY is hosted within the EMIS and TPP environments which are accredited to the ISO 27001 information security standard and are NHS IG Toolkit compliant^23^;

Patient data has been pseudonymised for analysis and linkage using industry standard cryptographic hashing techniques; all pseudonymised datasets transmitted for linkage onto OpenSAFELY are encrypted; access to the NHS England OpenSAFELY COVID-19 service is via a virtual private network (VPN) connection; the researchers hold contracts with NHS England and only access the platform to initiate database queries and statistical models; all database activity is logged; only aggregate statistical outputs leave the platform environment following best practice for anonymisation of results such as statistical disclosure control for low cell counts^24^.

The service adheres to the obligations of the UK General Data Protection Regulation (UK GDPR) and the Data Protection Act 2018. The service previously operated under notices initially issued in February 2020 by the the Secretary of State under Regulation 3(4) of the Health Service (Control of Patient Information) Regulations 2002 (COPI Regulations), which required organisations to process confidential patient information for COVID-19 purposes; this set aside the requirement for patient consent.^25^ As of 1 July 2023, the Secretary of State has requested that NHS England continue to operate the Service under the COVID-19 Directions 2020.^26^ In some cases of data sharing, the common law duty of confidence is met using, for example, patient consent or support from the Health Research Authority Confidentiality Advisory Group.^27^

Taken together, these provide the legal bases to link patient datasets using the service. GP practices, which provide access to the primary care data, are required to share relevant health information to support the public health response to the pandemic, and have been informed of how the service operates.

This study was approved by the Health Research Authority (REC reference 20/LO/0651) and by the LSHTM Ethics Board (reference 21863).

## Data access and verification

Access to the underlying identifiable and potentially re-identifiable pseudonymised electronic health record data is tightly governed by various legislative and regulatory frameworks, and restricted by best practice. The data in the NHS England OpenSAFELY COVID-19 service is drawn from General Practice data across England where EMIS and TPP are the data processors.

EMIS and TPP developers initiate an automated process to create pseudonymised records in the core OpenSAFELY database, which are copies of key structured data tables in the identifiable records. These pseudonymised records are linked onto key external data resources that have also been pseudonymised via SHA-512 one-way hashing of NHS numbers using a shared salt. University of Oxford, Bennett Institute for Applied Data Science developers and PIs, who hold contracts with NHS England, have access to the OpenSAFELY pseudonymised data tables to develop the OpenSAFELY tools.

These tools in turn enable researchers with OpenSAFELY data access agreements to write and execute code for data management and data analysis without direct access to the underlying raw pseudonymised patient data, and to review the outputs of this code. All code for the full data management pipeline - from raw data to completed results for this analysis - and for the OpenSAFELY platform as a whole is available for review at github.com/OpenSAFELY.

## Contributions

CDA is first author

BMK and SB are joint last - BMK responsible for research and clinical informatics input whilst SB led development and oversight of the OpenSAFELY platform.

Conceptualization: CDA, BMK, and CW. Data curation: CDA, LF, HJC, WJH, and AJW. Formal analysis: CDA. Funding acquisition: BG. Methodology: CDA, LF, and AJW. Project administration: CDA, BMK, and CW. Software: CDA, LF, HJC, WJH, AM, RS, TW, SD, PI, ID, SM, TOD, BBC, LB, CB, JC, SOH, DA, RJ, AJW, BG, BMK and SB. Supervision: BMK, and WJH. Validation: CDA, CW, BMK, HJC and WJH. Visualisation: CDA, CW, BMK, HJC and WJH. Writing - original draft: CDA. Writing - review & editing: CDA, CW, WJH, HJC and BMK.

## Box 1

CO@h and COPD005 SNOMED CT Codes

| CO@h SNOMED CT code |  |
| --- | --- |
| SNOMED CT CODE | Term |
| 1325251000000106 | Referral to telehealth pulse oximetry monitoring service (procedure) |
| 1325261000000109 | Referral by telehealth pulse oximetry monitoring service (procedure) |
| 1325271000000102 | Discharge from telehealth pulse oximetry monitoring service (procedure) |
| 1325201000000105 | Telehealth pulse oximetry monitoring ended (situation) |
| 1325191000000108 | Telehealth pulse oximetry monitoring started (situation) |
| 1325211000000107 | Provision of pulse oximeter (procedure) |
| 1325221000000101 | Telehealth pulse oximetry monitoring not appropriate (finding) |
| 1325241000000108 | Telehealth pulse oximetry monitoring declined (situation) |
| 1325281000000100 | Discussion about telehealth pulse oximetry monitoring (procedure) |
| 1325681000000102 | Has access to pulse oximeter (finding) |
| 1325701000000100 | Oxygen saturation at periphery equivocal (finding) |
| 1325691000000100 | Oxygen saturation at periphery unknown (finding) |
| COPD005 codes |  |
| SNOMED CT Code | Term |
| 252465000 | Pulse oximetry |
| 431314004 | Peripheral oxygen saturation |
| 866661000000106 | Peripheral blood oxygen saturation on room air at rest |
| 866701000000100 | Peripheral blood oxygen saturation on supplemental oxygen at rest |
| 927981000000106 | Baseline oxygen saturation at periphery |

## Box 2

Eligible Criteria (at the time of the study: October 2020 and July 2021)

NHS guidance criteria recommends the COVID Oximetry @home pathway should be available to people who are:

- diagnosed with COVID-19: either clinically or positive test result and
- symptomatic and either
  ∘ aged 65 years or older or
  ∘ Under 65 years and clinically extremely vulnerable (CEV) to coronavirus or where clinical judgement applies, taking into account multiple COVID risk factors.

We investigated this population in TPP only. OpenSAFELY is under rapid iteration and development and as a consequence, the features and data available in the underlying data environments differ.

All elements of the eligibility criteria are available in OpenSAFELY-TPP and are under development in OpenSAFELY-EMIS.

